# Effect of smoking cessation on CT imaging in patients with Chronic Obstructive Pulmonary Disease: A systematic review

**DOI:** 10.1101/2020.02.11.20022129

**Authors:** Daryl Cheng, Siddharth Agarwal, Joseph Jacob, John R Hurst

**Affiliations:** UCL Respiratory, University College London, London, UK; Department of Radiology, University of Nottingham, Nottingham, UK; Centre for Medical Image Computing, University College London, London, UK

## Abstract

**Background:** Smoking cessation is the only intervention known to affect disease progression in patients with COPD as measured by the rate of change in forced expiratory volume/1s (FEV_1_) over time. The need for new drugs to modify the progression of COPD is well recognised. We hypothesised that changes on CT in relation to smoking cessation may relate to changes in response to disease-modifying drugs, and therefore as a novel quantitative biomarker of drug efficacy. CT biomarkers of emphysema and airway wall thickness are increasingly used in research, but there has not been a systematic appraisal of the evidence to assess how these biomarkers evolve with a change in smoking exposure in COPD patients.

**Methods:** We searched MEDLINE, Embase, the Cochrane Library (Cochrane Database of Systematic Reviews, Cochrane Central Register of Controlled Trials (CENTRAL)), and Web of Science to 10^th^ September 2019. We included longitudinal studies of smoking COPD patients who had CT scans before and after smoking cessation. Two review authors (DC, SA) independently screened studies, extracted outcome data and assessed the risk of bias, with a third reviewer (JRH) arbitrating conflicts.

**Results:** Four studies were included in the final analysis. Three studies measured CT markers of lung density, which all, perhaps counter-intuitively, showed a significant decrease with smoking cessation. One study measured CT markers of airway wall thickness, which also significantly decreased with smoking cessation.

**Authors’ conclusions:** Smoking cessation in COPD patients causes a fall in lung density, but the magnitude of the effect has not been rigorously assessed. One study has reported a decrease in airway wall thickness with smoking cessation. The number of studies is small, with some risk of bias. This question remains important for COPD researchers and requires further studies, in particular to assess whether changes with smoking cessation may model changes in response to novel pharmaceutical agents, and how to handle change in smoking status in relation to longitudinal observational imaging studies in COPD.

## 1 Introduction

Chronic obstructive pulmonary disease (COPD) is a condition characterised by airflow limitation and persistent respiratory symptoms. It is the fourth leading cause of death worldwide^1^ and its prevalence is increasing^2^. Disease progression has classically been assessed by measures such as the rate of change in forced expiratory volume over 1 second (FEV_1_). Despite the development of drugs to reduce symptoms and prevent exacerbations in COPD, smoking cessation is the only proven way to slow the progression of disease ^3^, as measured by rate of decline in FEV1, and remains the only disease-modifying intervention we can offer patients. In part, this likely reflects the heterogeneity in rate of FEV1 decline between patients with COPD. There is currently much interest in developing disease-modifying interventions in early COPD and one limitation is a current lack of quantitative biomarkers of disease progression.

Since its introduction more than four decades ago, computed tomography (CT) is now widespread in clinical practice and research. It has unique strengths in terms of image resolution and speed of acquisition which mean that it has utility in a variety of pulmonary conditions. In COPD, quantitative regional assessment of emphysema is already used to guide use of endobronchial valves, and has been used as a biomarker of response to alpha-1 antitrypsin augmentation, whilst quantitative assessments of airway wall geometry and functional small airway dysfunction have proved useful phenotyping tools in COPD research.

These CT biomarkers hold further promise as biomarkers of treatment response in COPD. We hypothesised that quantitative imaging biomarkers that change in relation to smoking cessation – the archetypal disease modifying intervention in COPD – may also be useful as markers to treatment response in relation to novel drugs.

We have therefore conducted a systematic review of studies examining the CT changes which occur as a result of smoking cessation in COPD patients. We aimed to synthesise the range of quantitative biomarkers measured, and the direction and size of effect with intervention. We provide a robust quality assessment of our included studies. Our results will be of interest to those developing imaging biomarkers of treatment response in COPD.

Our review question was: how does CT imaging in patients with COPD change when they stop smoking compared to if they continue smoking? Our outcome measures are quantitative changes in CT scan appearances.

## 2 Methods

### 2.1 Protocol and registration

A review protocol was published on the 4^th^ September 2019 on PROSPERO [ref]. The protocol was structured according to the Preferred reporting items for systematic review and meta-analysis protocols (PRISMA-P) 2015 statement ^8^. The protocol registration number is CRD42019144555.

### 2.2 Criteria for review

#### 2.2.1 Types of study

We included original research studies which report the outcome (quantitative CT scan appearances) in our population (COPD patients) and which meet our definition of change in exposure (successful smoking cessation).

We included longitudinal studies, with no other restrictions on study design. Studies could be case controlled, cohort, or observational, prospective or retrospective. Since randomisation to continued smoking would be unethical, randomisation was not an inclusion criterion. Studies in any language were included.

We excluded studies which recorded outcome measures during acute hospitalization or during COPD exacerbations. We excluded review articles. We excluded longitudinal studies which reported outcome measures only before or only after intervention.

#### 2.2.2 Types of patient

Our target patients were adults with a diagnosis of COPD with a significant smoking history. We did not make restrictions as to the diagnostic criteria the study authors chose for COPD diagnosis. We did not make restrictions as to the definition of ‘significant smoking’.

#### 2.2.3 Types of intervention

Intervention was successful smoking cessation between two outcome time-points (CT scan). We did not place any restriction on the duration of smoking cessation. If a control group was used in a study, this would be taken as patients who continued to smoke until the second time point. A lack of a control group was not an exclusion criterion since some studies may elect to use extrapolation from longitudinal data as a comparison.

#### 2.2.4 Types of outcome and prioritisation

The main outcomes were: (1) change in quantitative CT biomarkers of emphysema; and (2) change in quantitative CT biomarkers of airway wall thickness.

We also report qualitative change in CT appearances.

### 2.3 Search methods for identification of studies

#### 2.3.1 Information sources

The bibliographic databases we searched included: OVID MEDLINE, EMBASE, The Cochrane Library (Cochrane Database of Systematic Reviews, Cochrane Central Register of Controlled Trials (CENTRAL)), and Web of Science. Other sources included articles identified through discussion with experts, including experts on the review team.

#### 2.3.2 Search strategy

The search strategy was formulated to include all articles which included the topics of chronic obstructive pulmonary disease, smoking cessation, and computed tomography. The technical differences in compiling this search were translated across the different bibliographic database. The Evidence Services Librarian at University College London was consulted to help build a robust search strategy during the protocol stage of the review. The search strategy was also appraised using the Peer Review of Electronic Search Strategies (PRESS) guideline ^9^. The full search strategy for each of the databases in included as appendix A. The search was carried out on 10^th^ September 2019 for all three databases.

### 2.4 Data collection and analysis

#### 2.4.1 Study selection

Titles of studies were retrieved using the search strategy from our information sources. These studies were combined with others found from other sources. Duplicates were removed. Two reviewers (DC and SA) independently screened the study titles for potential eligibility to meet the inclusion criteria. The abstracts for these studies were retrieved for a further round of screening. All studies which met inclusion screening by abstract had full text retrieved. The full texts of these articles were screened by two review members independently, and any discordance about eligibility were resolved by a third member of the review team (JRH).

#### 2.4.2 Data extraction and management

Data was extracted using a standardised form (6Appendix B). Data was extracted independently by two study authors and any discrepancies resolved through discussion.

#### 2.4.3 Assessment for risk of bias in individual studies

To assess for the risk of bias in individual studies, the formal risk of bias for internal validity tool ‘Risk Of Bias In Non-randomised Studies - of Interventions’ (ROBINS-I) was used ^10^. We first attempted to resolve disagreement between two reviewer judgements by consensus discussion; then by majority opinion with the third reviewer.

#### 2.4.4 Data synthesis

A data extraction table was produced using the data items outlined above. We have provided a narrative synthesis of the findings of the included studies and reported the direction and magnitude of the effect size of summary statistics. We have also summarised the homogeneity and perceived efficacy of the smoking cessation intervention performed, and the different CT metrics reported by each study.

We have not proceeded to any formal meta-analysis. This is because of the small number of included studies, the heterogenous CT metrics used, the difference in time between stopping smoking, and the heterogenous clinical characteristics of the patient populations.

## 3 Results

### 3.1 Results of search

A total of 1467 studies were retrieved from the search for screening and combined with one study collected from discussion with experts. 194 duplicates were removed to give a total of 1273 studies to screen by title and abstract. At title and abstract screening, reviewer DC included 12 studies, whereas SA included 14. 6 of these were in conflict, with a total of 16 studies included by a minimum of 1 reviewer. These studies had full text retrieved for screening, of which 5 were included in the final analysis. Study selection is summarised in the PRISMA flowchart (Figure 1).

#### 3.2 Excluded studies

1257 studies were excluded by screening by title and abstract. 11 studies were excluded by full text, and the reasons for this are detailed in table 5.4 ‘Characteristics of excluded studies‘. The most common reason was that there was no evidence of smoking cessation between the two time points, and studies examined CT differences between former smokers and current smokers (n=5). The next most common reason was that the population investigated were not COPD patients (n=4).

#### 3.3 Included studies

The five remaining studies were published between 2011 and 2019. Details of the included studies are included in the table 5.3 ‘Characteristics of included studies‘. Four studies were published in English. One study was published in Chinese and was translated by reviewer DC for the review team.

#### 3.4 Risk of bias

##### 3.4.1 Risk of bias across studies

The formal risk of bias for internal validity tool ‘Risk Of Bias In Non-randomised Studies – of Interventions’ (ROBINS-I) was used to assess biases across all included studies. The confounders selected for consideration included: socioeconomic differences between intervention groups; and severity of disease and differences in previous history of smoking between intervention groups. Socioeconomic factors may influence co-variates such as environmental exposures which could influence the outcome. Patients with severe disease or a stronger smoking history may be less likely to successfully quit smoking and thus baseline lung pathology could influence the outcome.

Co-interventions such as nicotine replacement therapy or exercise were possibly different between intervention groups, but this was not thought likely to impact outcomes.

The ‘ideal’ randomised trial design was an individually randomised trial of COPD patients to either continue smoking or stop smoking immediately after the first outcome measure and remain abstinent until a repeat outcome measure at 1 year.

##### 3.4.2 Risk of bias within included studies

The ROBINS-I tool was used to evaluate individual studies for potential bias due to confounding, participant selection, intervention classification, deviation from intervention, missing data, outcome measurement, and selective reporting. The authors’ risk of bias judgements for the individual studies are included in the supplementary table 5.3 ‘Characteristics of included studies‘.

Due to a critical risk of bias in the selective reporting, of the 5 included studies, one^11^ has not been included in the narrative synthesis, and the results presented hereafter are from the remaining four studies. The excluded study compared the change in %LAA-950 between the 5 lobes of the lung between two time points without evidence of multi-comparison correction for statistical significance.

The included studies detailed numbers of patients in each cohort who successfully stopped smoking according to their definition of smoking cessation. These definitions differed however, making direct comparison difficult. For example, the monitoring differed, with some authors replying on smoking diaries, and some on self-reported questionnaires. One study used carbon monoxide monitoring. None of the studies reported the time duration from smoking cessation to the outcome measures (CT scan).

#### 3.5 Main outcomes

##### 3.5.1 Quantitative change in CT markers of emphysema

Two of the included studies involving 81 patients who stopped smoking, reported changes in quantitative CT markers of lung density using the percentage or relative area of lung below an attenuation threshold ^12,13^. Both studies reported a significant increase in the %LAA/RA-950 or -910 HU, representing a fall in lung density. Both also reported a significant decrease in the 15^th^ percentile of lung density (PD15), again representing a fall in lung density with smoking cessation.

Shaker and Hlaing demonstrated that up to a year after smoking cessation, there is a significant decrease in lung density (n=36, n=45 respectively), a difference that was demonstrated to be greater than a continued smoking control group from Shaker’s original study (Table 5.2).

Hlaing *et al* (2015) also demonstrated a significant reduction in the mean lung density (MLD, -7.7HU, SD=2.3, p<0.001).

##### 3.5.2 Quantitative change in CT markers of airway wall thickness

Only one study involving 203 patients who stopped smoking reported changes in airway wall metrics on CT with smoking cessation ^14^. The authors reported a significant decrease in wall thickness of - 0.18mm (95% CI -0.23 to -0.13, p<0.001) on smoking cessation between two scans five years apart. The authors did not comment on duration of smoking cessation or time from smoking cessation to second scan but defined smoking cessation as self-reported abstinence from smoking after the first scan.

#### 3.6 Other outcomes

##### 3.6.1 Qualitative change in CT appearances

One study reported qualitative changes in appearances of emphysema on smoking cessation. This was performed blinded, with the radiologist comparing scans for emphysema, ground glass opacification or micronodules, documenting whether these features were more, less, or the same quantity as the baseline scan. The study reported no significant difference in the presence of emphysema visually with smoking cessation but did report a decrease in the presence of micronodules.

##### 3.6.2 Change in FEV_1_ with smoking cessation

Three of the five studies reported a change in FEV_1_ with smoking cessation. Two papers reported no significant difference in this measure at one year, whilst Hlaing *et a*l (2015) reported a significant decline in FEV_1_ after 1 year sustained smoking abstinence (-33ml, p<0.001). The authors noted that this change was less than patients who continued to smoke.

Dhariwal *et al* (2014) reported a transient improvement in FEV_1_ of mean 184ml at 6 weeks, but this decreased to 81ml at 12 weeks and not fully maintained at 1 year. Shaker *et al* (2011) reported no significant change in FEV_1_ within one year of smoking cessation (72ml, SD=47ml, p=0.14).

### 4 Discussion

#### 4.1 Summary of main results

##### 4.1.1 Measures of Lung Density

In summary, quantitative CT metrics of lung density fall with smoking cessation. The direction of effect of three markers across the four studies included was consistent. Relative area of low attenuation (RAA) - RAA-910 and RAA-950 are defined as the percentage of lung pixels, or the relative area, with lower than 910 and 950 Hounsfield units respectively (approximately the density of air). The advantage of these measures and the mean lung density is that they are objective, replicable and are quick to assess using automated software, however may be underestimated in the presence of transient consolidation. MLD suffers from the additional disadvantage in that it includes non-airspace densities even when the lungs are segmented meticulously. Density of the 15th percentile (PD15) is similarly an automated density measure, but one that is not affected by ‘outliers’ e.g. consolidation.

##### 4.1.2 Measures of Airway Thickness

Only one study reported the change in airway wall thickness, suggesting that this decreases with smoking cessation, but because this result has not been replicated, and difficult to interpret with confidence. The Pi10 measure, the square root wall thickness for all airways with an internal diameter of 10mm, is a global measure of wall thickness using automated software. This measure is objective and replicable, but importantly is different to measures of lung density and therefore is theoretically prone to a different set of confounders. Charbonnier [**] demonstrated that the same cohort of quitters had both a decrease in Pi10 and RAA-910, suggesting that this effect is not due to a different patient cohort.

#### 4.2 Limitations

Overall, a small number of studies were available for evidence synthesis, and they varied in quality and risk of bias. Of the five studies included after the final search, one had a critical risk of bias. The studies addressed prior smoking history and severity of COPD, with varying degrees of rigour.

At the outcome level, most of the studies used differing outcome measures which made direct comparison difficult and meta-analysis impossible. The outcomes chosen were also limited by the scans performed, with newer biomarkers such as parametric response mapping (PRM) not yet evaluated. The studies also varied in terms of length of follow up.

At the review level, limitations were the small number of studies which met our final inclusion criteria. Large longitudinal cohorts such as ECLIPSE likely contain the data required for this analysis, but researchers have not analysed the effect of smoking cessation ^16^ There likely exists a sizable dataset of imaging data for in lung cancer screening populations, and we were careful not to exclude these from our search, but we did not identify any which examined the subset of COPD patients. Even within the broader question of the longitudinal anatomical evolution of COPD is not well studied. As noted by Coxson et al, cohorts were limited by their lack of specific focus on COPD patients.

#### 4.3 Conclusions

CT measures of COPD equate to lower lung density and thickened airways, although these studies demonstrate this more in the smoking cessation groups.

Pathologies that affect airway wall thickness and lung density can confound these CT biomarkers. Smoking may promote an inflammatory state which increases the density of lung parenchyma and thickens the airway walls^15^. The implication is that these biomarkers may be largely measuring transient inflammation, and that the hypothesised mechanism of lower lung density and thickened airways – which could be interpreted as COPD progression – requires further study. It is worth noting that no study scanned beyond 5 years.

The pulmonary CT imaging changes that occur with smoking cessation in patients with COPD needs further study, and it is not at present possible to test the hypothesis that smoking-cessation changes on CT in COPD may be useful as biomarkers of drug intervention.

### 5. Figures and tables

#### 5.1 Figure 1: PRISMA diagram

**Figure 1.**
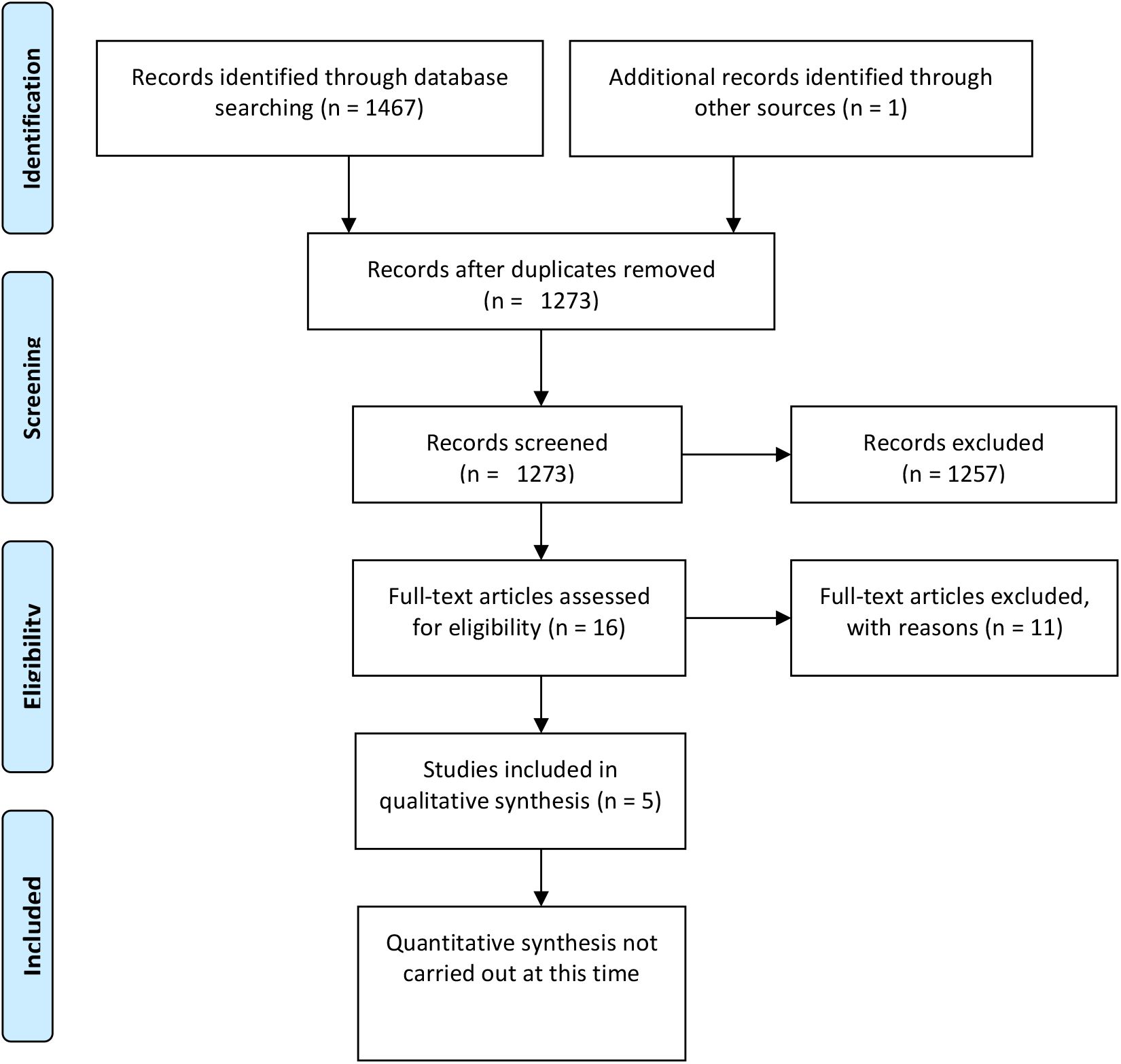
PRISMA diagram illustrating numbers of studies included ^8^.

### 5.2

**Table 1.**
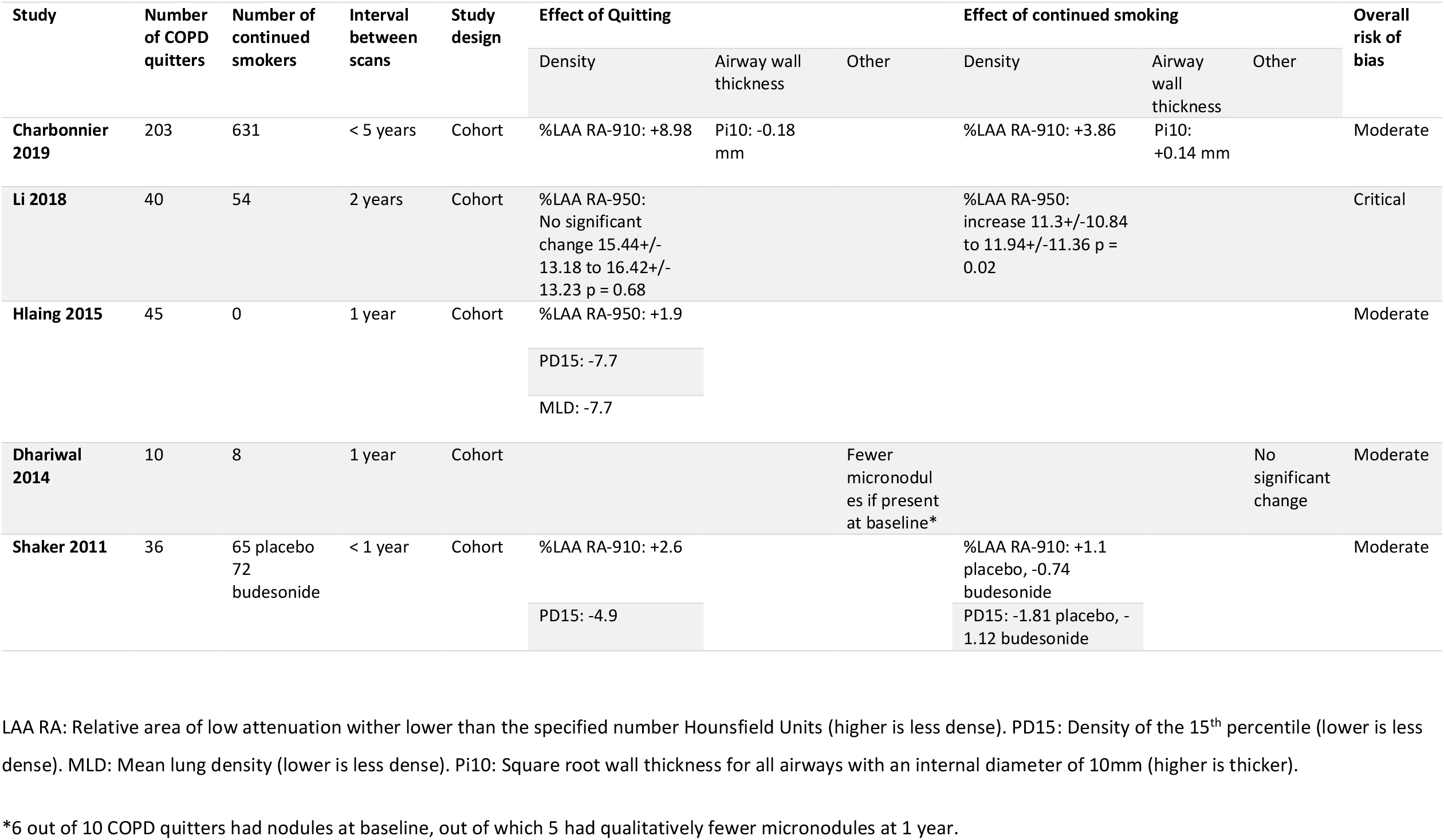

### 5.3 Characteristics of included studies

#### 5.3.1 Charbonnier et al 2019

|  |  |  |
| --- | --- | --- |
| Methods | Secondary analysis on a cohort recruited to study longitudinal history of COPD in the United States. Study data spanned 5 years. Analysis performed on inspiration only CT using automated tools. Unclear if smoking cessation interventions were administered. |  |
| Participants | Patients with COPD and matched smoking controls. In this analysis, Case N = 203, Control N = 631. Female = 47.6%. Mean age = 55.8 (7.4). Mean Pack year history = 42 (22.3) |  |
| Interventions | The intervention group were defined as patients who stopped smoking after visit 1. The control group were defined as patients who continued to smoke. |  |
| Outcomes | Change in Pi10 |  |
| Notes |  |  |
| Risk of bias |  |  |
| Bias | Authors' judgement | Support for judgement |
| Confounding | Low | Multivariate models adjusted for gender, age, BMI, pack years, TLC, BDR, smoking status, and LAA%-950. Analysis repeated per GOLD stage |
| Selection | Moderate | Unknown if start of follow-up and start of intervention coincide for most participants |
| Classification | Moderate | Unclear if all patients in smoking cessation analysis were in COPD cohort or if included smoking controls |
| Deviation | Moderate | Relied on self-reported smoking cessation |
| Missing data | Low | No participants excluded on basis of missing data |
| Measurement | Low | Use of automated tools only |
| Reporting | Low | Subgroup and multiple comparisons avoided |

#### 5.3.2 Li et al 2018

This study was not included in the narrative synthesis due to critical risk of bias.

|  |  |  |
| --- | --- | --- |
| Methods | Retrospective cohort analysis chest CT images of patients with COPD at two time points, with a two-year interval. Subgroup analyses of differences in automated CT metrics of lung density by smoking cessation |  |
| Participants | 130 patients identified meeting inclusion criteria of COPD patients with appropriate CT imaging between 40-80 years old and without concomitant lung disease. |  |
| Interventions | Retrospectively reported smoking cessation on hospital database records, unknown method of reporting, unknown measurement of deviation from intervention |  |
| Outcomes | %LAA-950, PD15 |  |
| Notes |  |  |
| <u>Risk of bias</u> | Overall risk of bias - Critical |  |
| Bias | Authors' judgement | Support for judgement |
| Confounding | Moderate | Start of follow-up and start of intervention does not coincide for most participants |
| Selection | Serious | Selection of participants into the analysis based on participant characteristics observed after the start of intervention |
| Classification | Moderate | Information used to define intervention groups not recorded at start of intervention |
| Deviation | Moderate | No evidence of monitoring of adherence to intervention |
| Missing data | Moderate | Participants were excluded due to missing data on other variables needed for the analysis |
| Measurement | Moderate | Unclear if measurement was blinded |
| Reporting | Critical | The reported effect estimate was likely to be selected, based on the results, from multiple outcome *measurements* within the outcome domain, as well as different *subgroups* |

#### 5.3.3 Hlaing et al 2015

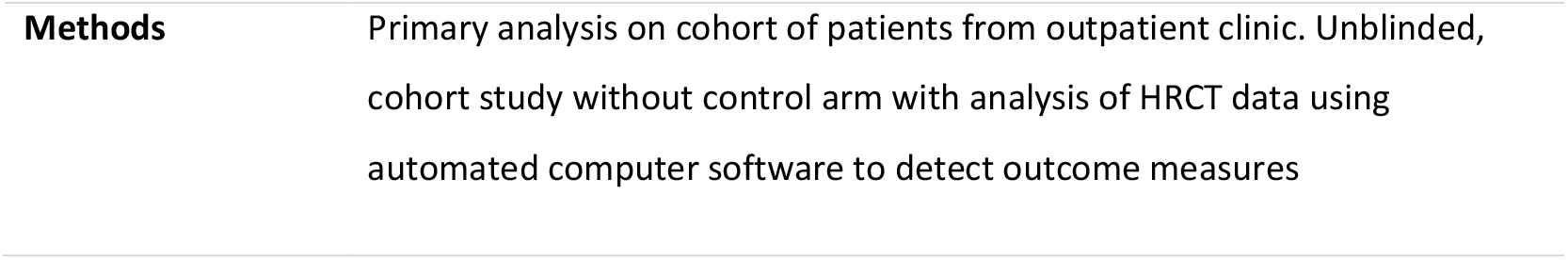
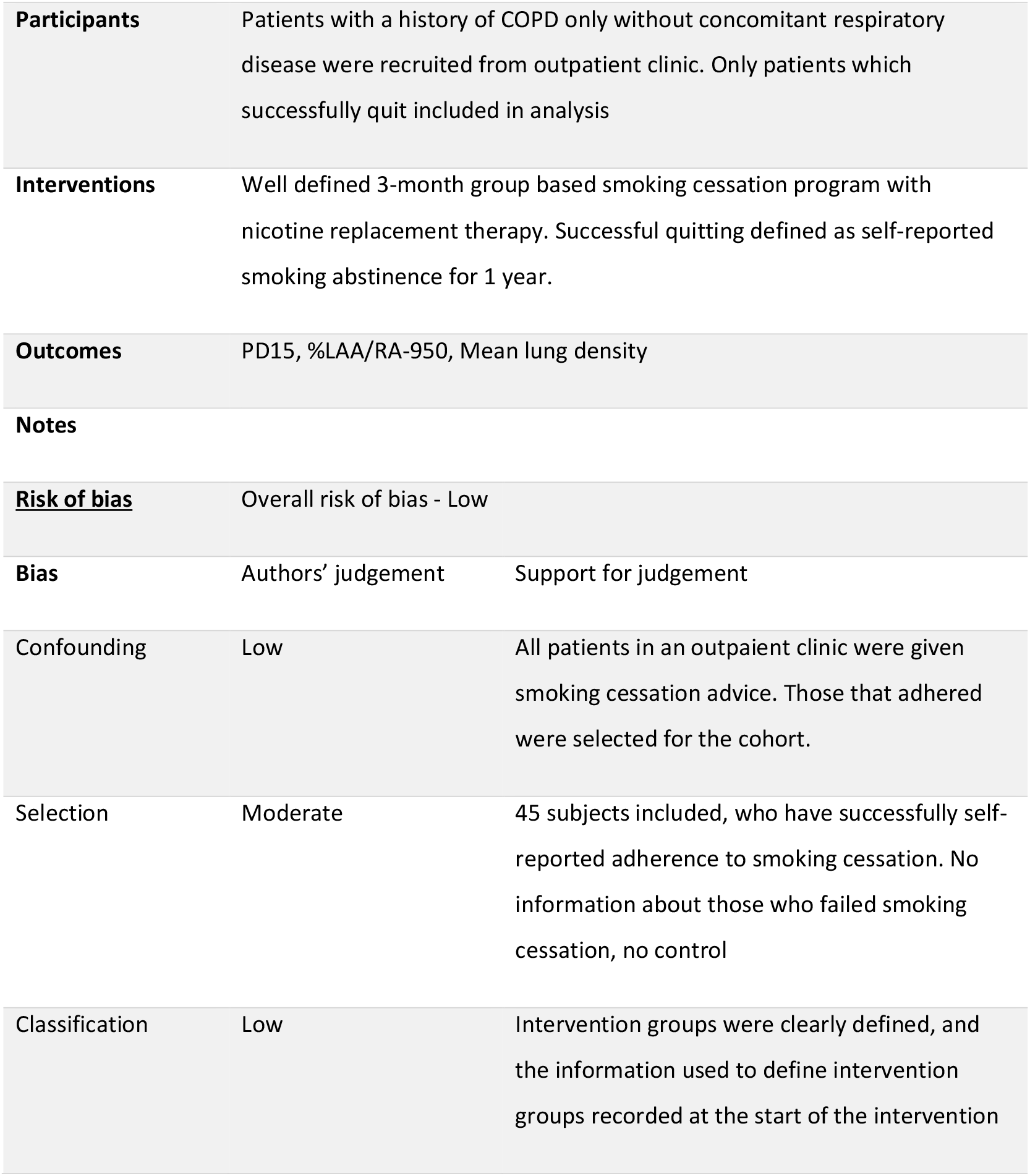
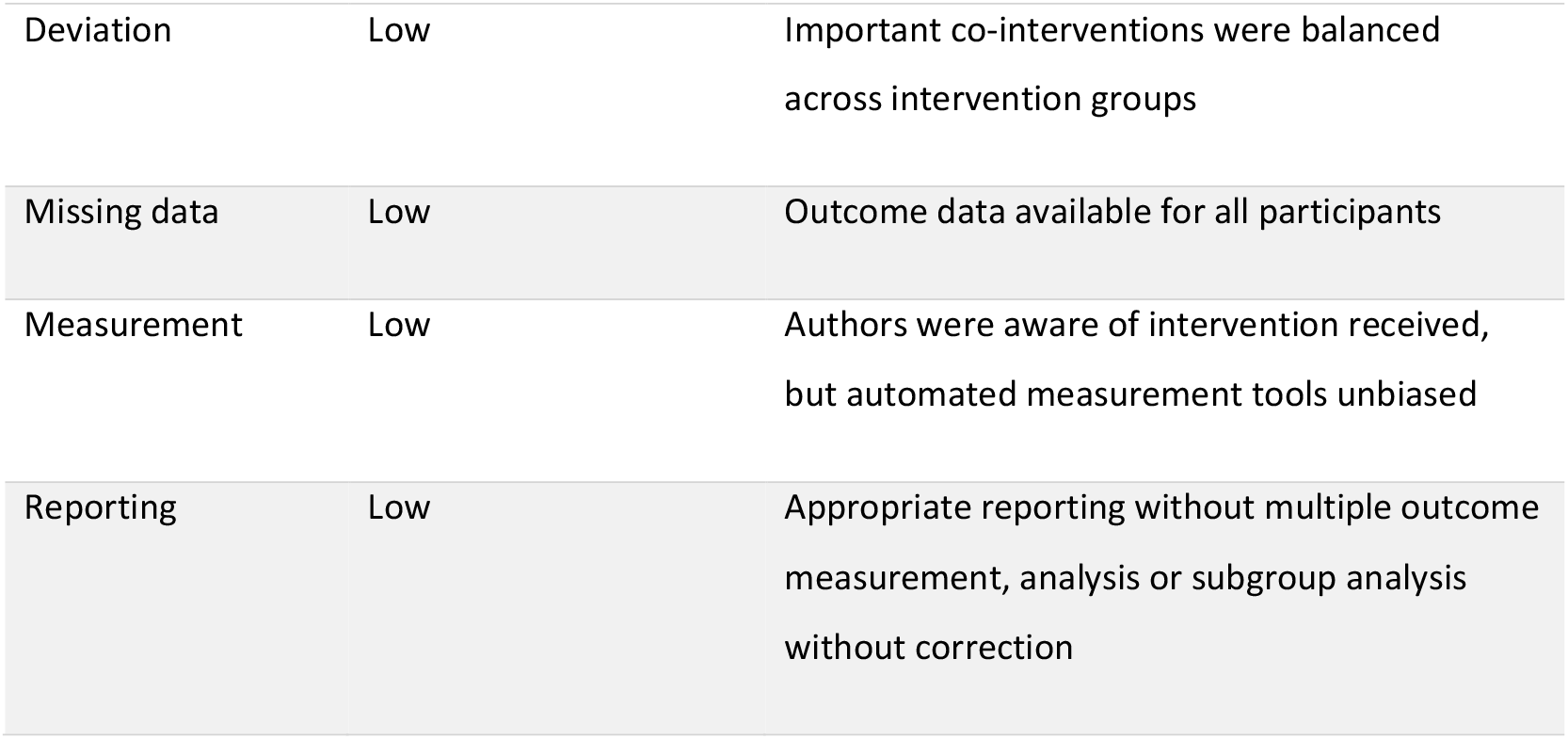

|  |  |
| --- | --- |
| **Methods** | Primary analysis on cohort of patients from outpatient clinic. Unblinded, cohort study without control arm with analysis of HRCT data using automated computer software to detect outcome measures |

|  |  |  |
| --- | --- | --- |
| Participants | Patients with a history of COPD only without concomitant respiratory disease were recruited from outpatient clinic. Only patients which successfully quit included in analysis |  |
| Interventions | Well defined 3-month group based smoking cessation program with nicotine replacement therapy. Successful quitting defined as self-reported smoking abstinence for 1 year. |  |
| Outcomes | PD15, %LAA/RA-950, Mean lung density |  |
| Notes |  |  |
| <u>Risk of bias</u> | Overall risk of bias - Low |  |
| Bias | Authors' judgement | Support for judgement |
| Confounding | Low | All patients in an outpatient clinic were given smoking cessation advice. Those that adhered were selected for the cohort. |
| Selection | Moderate | 45 subjects included, who have successfully self-reported adherence to smoking cessation. No information about those who failed smoking cessation, no control |
| Classification | Low | Intervention groups were clearly defined, and the information used to define intervention groups recorded at the start of the intervention |
| Deviation | Low | Important co-interventions were balanced across intervention groups |
| Missing data | Low | Outcome data available for all participants |
| Measurement | Low | Authors were aware of intervention received, but automated measurement tools unbiased |
| Reporting | Low | Appropriate reporting without multiple outcome measurement, analysis or subgroup analysis without correction |

#### 5.3.4 Dhariwal et al 2014

|  |  |
| --- | --- |
| **Methods** | Primary analysis of cohort of patients recruited for HRCT data. Subjects meeting inclusion criteria were screened for spirometric confirmation of COPD. HRCTs performed prior to smoking cessation, at week 12 and at week 52 of continued smoking abstinence. |
| **Participants** | Subjects recruited through smoking cessation clinics, newspaper advertising, local GPs and telephone helplines. |
| **Interventions** | All subjects offered either 6-week smoking cessation course of one-to-one counselling, with the option of nicotine replacement therapy |
| **Outcomes** | Qualitative reporting of emphysema and micronodules on CT data |
| **Notes** |  |
| **<u>Risk of bias</u>** |  |

| Bias | Authors' judgement | Support for judgement |
| --- | --- | --- |
| Confounding | Moderate | Prior smoking history not controlled for but otherwise subjects matched for |
| Selection | Low |  |
| Classification | Low |  |
| Deviation | Low | CO monitoring used to assess ongoing cessation |
| Missing data | Low | Outcome data available for all participants |
| Measurement | Low | Qualitative assessment by radiologists that were blinded to the intervention groups |
| Reporting | Moderate | Multiple subgroup analyses conducted although reported result is not selected from this |

#### 5.3.5 Shaker et al 2011

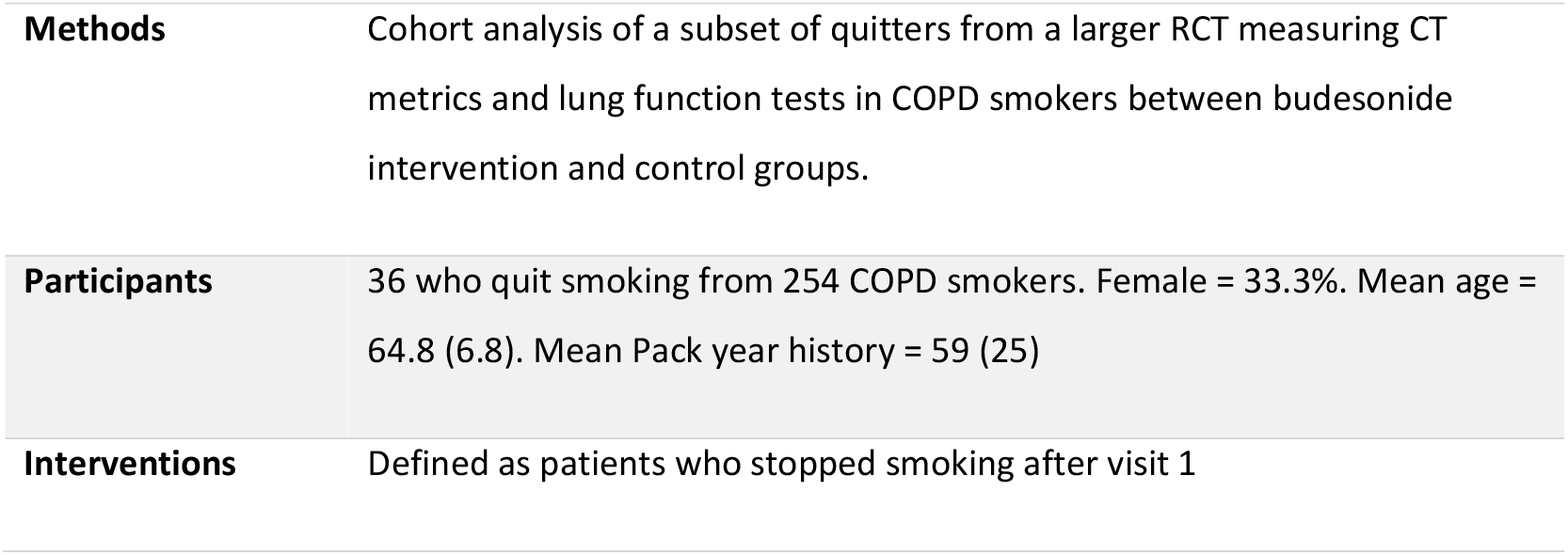
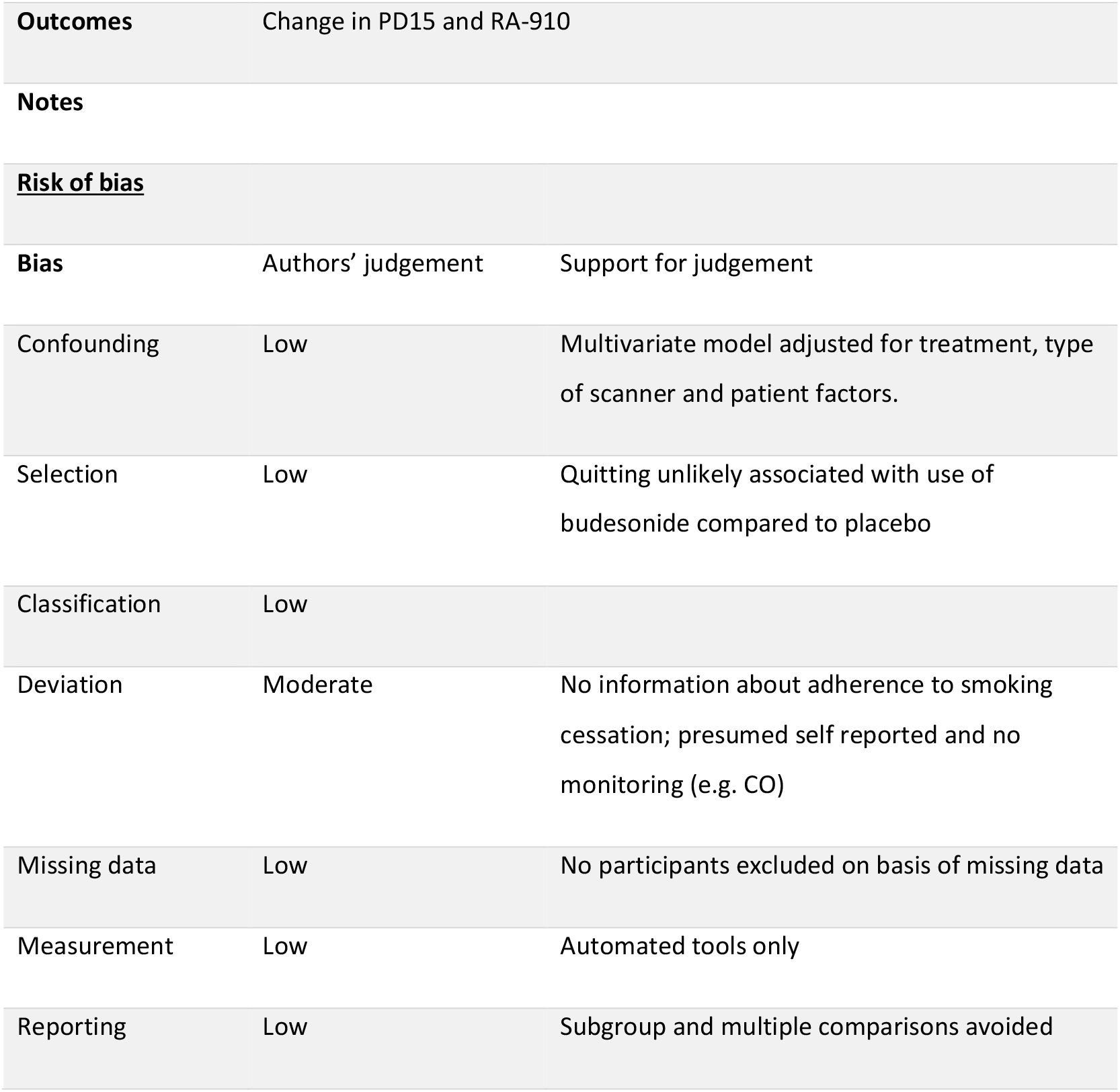

|  |  |
| --- | --- |
| **Methods** | Cohort analysis of a subset of quitters from a larger RCT measuring CT metrics and lung function tests in COPD smokers between budesonide intervention and control groups. |
| **Participants** | 36 who quit smoking from 254 COPD smokers. Female = 33.3%. Mean age = 64.8 (6.8). Mean Pack year history = 59 (25) |
| **Interventions** | Defined as patients who stopped smoking after visit 1 |

|  |  |  |
| --- | --- | --- |
| **Outcomes** | Change in PD15 and RA-910 |  |
| **Notes** |  |  |
| **<u>Risk of bias</u>** |  |  |
| **Bias** | Authors' judgement | Support for judgement |
| Confounding | Low | Multivariate model adjusted for treatment, type of scanner and patient factors. |
| Selection | Low | Quitting unlikely associated with use of budesonide compared to placebo |
| Classification | Low |  |
| Deviation | Moderate | No information about adherence to smoking cessation; presumed self reported and no monitoring (e.g. CO) |
| Missing data | Low | No participants excluded on basis of missing data |
| Measurement | Low | Automated tools only |
| Reporting | Low | Subgroup and multiple comparisons avoided |

#### 5.4 Characteristics of excluded studies

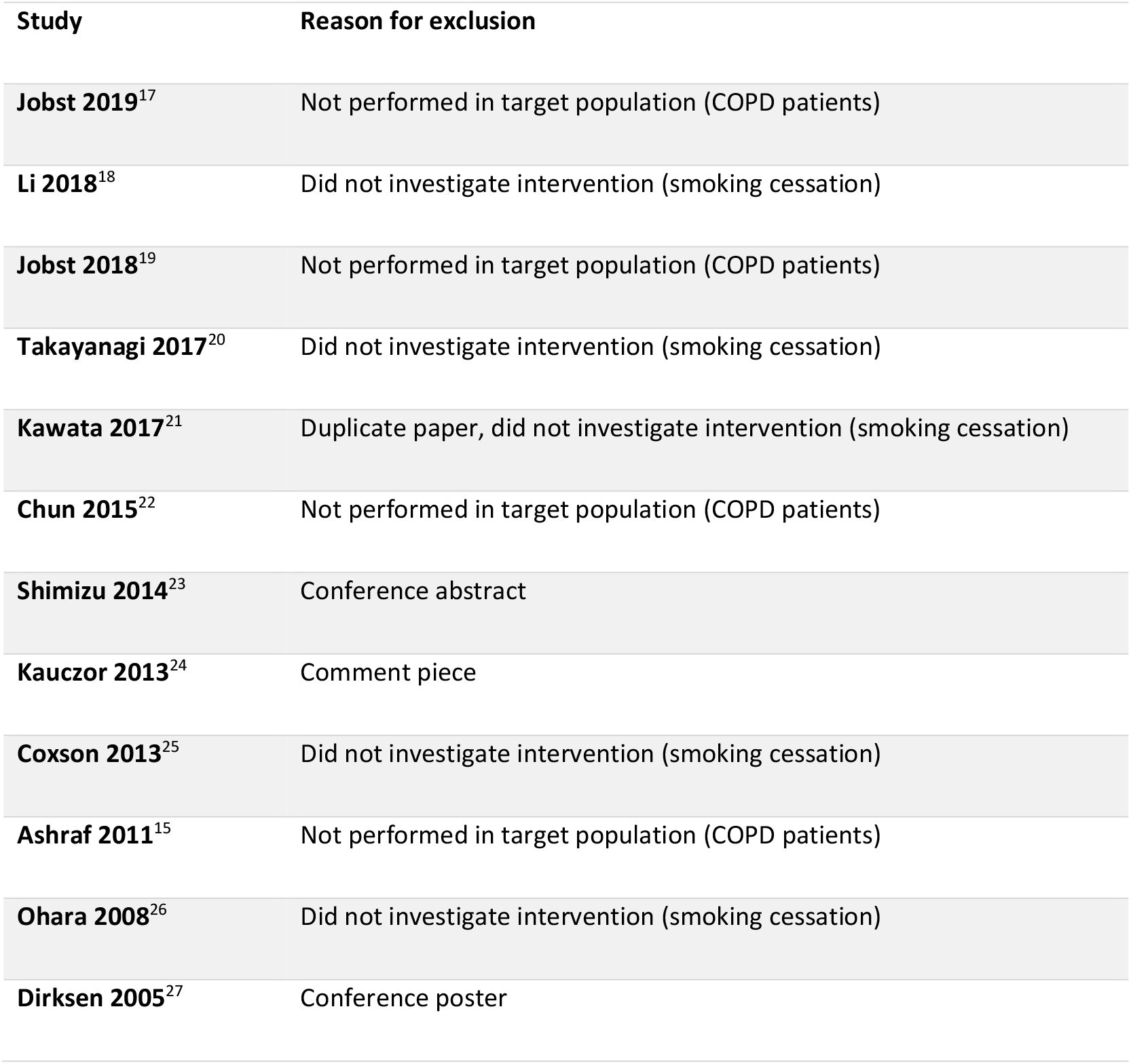

| Study | Reason for exclusion |
| --- | --- |
| **Jobst 2019**<sup>17</sup> | Not performed in target population (COPD patients) |
| **Li 2018**<sup>18</sup> | Did not investigate intervention (smoking cessation) |
| **Jobst 2018**<sup>19</sup> | Not performed in target population (COPD patients) |
| **Takayanagi 2017**<sup>20</sup> | Did not investigate intervention (smoking cessation) |
| **Kawata 2017**<sup>21</sup> | Duplicate paper, did not investigate intervention (smoking cessation) |
| **Chun 2015**<sup>22</sup> | Not performed in target population (COPD patients) |
| **Shimizu 2014**<sup>23</sup> | Conference abstract |
| **Kauczor 2013**<sup>24</sup> | Comment piece |
| **Coxson 2013**<sup>25</sup> | Did not investigate intervention (smoking cessation) |
| **Ashraf 2011**<sup>15</sup> | Not performed in target population (COPD patients) |
| **Ohara 2008**<sup>26</sup> | Did not investigate intervention (smoking cessation) |
| **Dirksen 2005**<sup>27</sup> | Conference poster |

## Data Availability

The data referred to in the manuscript is included either in the manuscript or the supplementary data at the end. The raw data is also available on request from the study authors.

## Appendix A Search Strategies

### A.1 Ovid MEDLINE® (1946 to August Week 5 2019)

1. exp Pulmonary Disease, Chronic Obstructive/ (52490)
2. Lung Diseases, Obstructive/ (18135)
3. (obstruct$ adj3 (pulmonary or lung$ or airway$ or airflow$ or bronch$ or respirat$)).mp. [mp=title, abstract, original title, name of substance word, subject heading word, floating sub-heading word, keyword heading word, organism supplementary concept word, protocol supplementary concept word, rare disease supplementary concept word, unique identifier, synonyms] (102899)
4. COPD.mp. [mp=title, abstract, original title, name of substance word, subject heading word, floating sub-heading word, keyword heading word, organism supplementary concept word, protocol supplementary concept word, rare disease supplementary concept word, unique identifier, synonyms] (36917)
5. COAD.mp. [mp=title, abstract, original title, name of substance word, subject heading word, floating sub-heading word, keyword heading word, organism supplementary concept word, protocol supplementary concept word, rare disease supplementary concept word, unique identifier, synonyms] (244)
6. emphysema*.mp. [mp=title, abstract, original title, name of substance word, subject heading word, floating sub-heading word, keyword heading word, organism supplementary concept word, protocol supplementary concept word, rare disease supplementary concept word, unique identifier, synonyms] (32495)
7. (chronic* adj3 bronchiti*).mp. [mp=title, abstract, original title, name of substance word, subject heading word, floating sub-heading word, keyword heading word, organism supplementary concept word, protocol supplementary concept word, rare disease supplementary concept word, unique identifier, synonyms] (10836)
8. 1 or 2 or 3 or 4 or 5 or 6 or 7 (138142)
9. smoking cessation/ or smoking reduction/ or smoking/ or exp pipe smoking/ or exp tobacco smoking/ (150642)
10. ((quit* or stop* or ceas* or cessation or abstain* or abstinen*) adj3 (smok* or tobacco or cigar*)).mp. [mp=title, abstract, original title, name of substance word, subject heading word, floating sub-heading word, keyword heading word, organism supplementary concept word, protocol supplementary concept word, rare disease supplementary concept word, unique identifier, synonyms] (41160)
11. 9 or 10 (157596)
12. exp Tomography, X-Ray Computed/ (410855)
13. CT scan.mp. [mp=title, abstract, original title, name of substance word, subject heading word, floating sub-heading word, keyword heading word, organism supplementary concept word, protocol supplementary concept word, rare disease supplementary concept word, unique identifier, synonyms] (44507)
14. CAT scan.mp. [mp=title, abstract, original title, name of substance word, subject heading word, floating sub-heading word, keyword heading word, organism supplementary concept word, protocol supplementary concept word, rare disease supplementary concept word, unique identifier, synonyms] (820)
15. CATSCAN.mp. [mp=title, abstract, original title, name of substance word, subject heading word, floating sub-heading word, keyword heading word, organism supplementary concept word, protocol supplementary concept word, rare disease supplementary concept word, unique identifier, synonyms] (3)
16. (comput* adj2 tomograph*).mp. [mp=title, abstract, original title, name of substance word, subject heading word, floating sub-heading word, keyword heading word, organism supplementary concept word, protocol supplementary concept word, rare disease supplementary concept word, unique identifier, synonyms] (300434)
17. 12 or 13 or 14 or 15 or 16 (546904)
18. 8 and 11 and 17 (769)

### A.2 EMBASE (Embase Classic & Embase 1947 to 2019 September 09)

1. exp chronic obstructive lung disease/ (125489)
2. COPD.mp. [mp=title, abstract, heading word, drug trade name, original title, device manufacturer, drug manufacturer, device trade name, keyword, floating subheading word, candidate term word] (82778)
3. (obstruct$ adj3 (pulmonary or lung$ or airway$ or airflow$ or bronch$ or respirat$)).mp. [mp=title, abstract, heading word, drug trade name, original title, device manufacturer, drug manufacturer, device trade name, keyword, floating subheading word, candidate term word] (201771)
4. COAD.mp. [mp=title, abstract, heading word, drug trade name, original title, device manufacturer, drug manufacturer, device trade name, keyword, floating subheading word, candidate term word] (455)
5. (chronic$ adj3 bronchiti$).mp. [mp=title, abstract, heading word, drug trade name, original title, device manufacturer, drug manufacturer, device trade name, keyword, floating subheading word, candidate term word] (22805)
6. emphysema*.mp. [mp=title, abstract, heading word, drug trade name, original title, device manufacturer, drug manufacturer, device trade name, keyword, floating subheading word, candidate term word] (56796)
7. 1 or 2 or 3 or 4 or 5 or 6 (271226)
8. exp smoking cessation/ (56555)
9. exp smoking reduction/ (149)
10. smoking cessation program/ (3217)
11. ((smok* or tobacco or cigar*) adj3 (quit* or stop* or ceas* or cessation or abstain* or abstinen*)).mp. [mp=title, abstract, heading word, drug trade name, original title, device manufacturer, drug manufacturer, device trade name, keyword, floating subheading word, candidate term word] (71196)
12. 8 or 9 or 10 or 11 (71244)
13. exp computer assisted tomography/ (1002372)
14. CT scan.mp. [mp=title, abstract, heading word, drug trade name, original title, device manufacturer, drug manufacturer, device trade name, keyword, floating subheading word, candidate term word] (98165)
15. CAT scan.mp. [mp=title, abstract, heading word, drug trade name, original title, device manufacturer, drug manufacturer, device trade name, keyword, floating subheading word, candidate term word] (1360)
16. CATSCAN.mp. [mp=title, abstract, heading word, drug trade name, original title, device manufacturer, drug manufacturer, device trade name, keyword, floating subheading word, candidate term word] (17)
17. (comput* adj2 tomograph*).mp. [mp=title, abstract, heading word, drug trade name, original title, device manufacturer, drug manufacturer, device trade name, keyword, floating subheading word, candidate term word] (992850)
18. 13 or 14 or 15 or 16 or 17 (1100405)
19. 7 and 12 and 18 (466)

### A.3 The Cochrane Library

1. MeSH descriptor: [Pulmonary Disease, Chronic Obstructive] explode all trees-4858
2. MeSH descriptor: [Lung Diseases, Obstructive] this term only-2522
3. COPD OR (chronic obstructive pulmonary disease*)-18567
4. COAD OR (chronic obstructive airway* disease*)-8305
5. (obstruct* NEAR/2 (pulmonary OR lung* OR airway* OR airflow* OR bronch* OR respirat*))-18034
6. chronic bronchitis-2597
7. emphysema*-1646
8. {OR #1-#7}-25813
9. MeSH descriptor: [Smoking Cessation] explode all trees-3779
10. MeSH descriptor: [Tobacco Use Cessation] explode all trees-94
11. MeSH descriptor: [Smoking Reduction] explode all trees-9
12. stop* smok*-2339
13. (“tobacco control intervention”):kw-0
14. (“smoking cessation treatment”):kw-108
15. smok* cessation-10210
16. (quit* OR stop* OR cessation OR ceas*) NEAR (smok* OR tobacco OR cigar*)-11130 17.
17. {OR #9-#16}-11930
18. MeSH descriptor: [Tomography, X-Ray] explode all trees-4975
19. CT scan*-10513
20. comput* NEAR/2 tomograph*-19483
21. CAT scan*-226
22. CATSCAN*-17

### A.4 Web of Science (All years 1900-2019)

1. TS=(COPD OR chronic obstructive pulmonary disease* OR COAD OR chronic obstructive airway* disease* OR chronic obstructive lung disease OR emphysema OR chronic bronchitis) [105,681]
2. TS=(CT OR comput* tomograph*OR CAT OR CATSCAN) [401,167]
3. TS=((smok* OR tobacco OR cigar*) AND (quit* OR stop* OR cessation OR ceas* OR giv* OR prevent* OR abstain* or abstinen*)) [97,885]
4. #3 AND #2 AND #1 [163]

## Appendix B Data extraction form

Included as supplementary figure

